# Bone canonical Wnt signaling is downregulated in type 2 diabetes and associates with higher Advanced Glycation End-products (AGEs) content and reduced bone strength

**DOI:** 10.1101/2023.10.06.23296647

**Authors:** Giulia Leanza, Francesca Cannata, Malak Faraj, Claudio Pedone, Viola Viola, Flavia Tramontana, Niccolò Pellegrini, Gianluca Vadalà, Alessandra Piccoli, Rocky Strollo, Francesca Zalfa, Alec Beeve, Erica L Scheller, Simon Tang, Roberto Civitelli, Mauro Maccarrone, Rocco Papalia, Nicola Napoli

## Abstract

Type 2 diabetes (T2D) is associated with higher fracture risk, despite normal or high bone mineral density. We reported that bone formation genes (*SOST* and *RUNX2*) and Advanced Glycation End-products (AGEs) were impaired in T2D. We investigated Wnt signaling regulation and its association with AGEs accumulation and bone strength in T2D from bone tissue of 15 T2D and 21 non-diabetic postmenopausal women undergoing hip arthroplasty. Bone histomorphometry revealed a trend of low mineralized volume in T2D [(T2D 0.249% (0.156-0.366) vs non-diabetic subjects 0.352% (0.269-0.454); p=0.053)], as well as reduced bone strength [T2D 21.60 MPa (13.46-30.10) vs non-diabetic subjects 76.24 MPa (26.81-132.9); p=0.002]. We also showed that gene expression of Wnt agonists *LEF-1* (p=0.0136) and *WNT10B* (p=0.0302) were lower in T2D. Conversely, gene expression of *WNT5A* (p=0.0232), *SOST* (p<0.0001) and *GSK3B* (p=0.0456) were higher, while collagen (*COL1A1*) was lower in T2D (p=0.0482). AGEs content was associated with *SOST* and *WNT5A* (r=0.9231, p<0.0001; r=0.6751, p=0.0322), but inversely correlated with *LEF-1* and *COL1A1* (r= -0,7500, p=0.0255; r= -0,9762, p=0.0004). *SOST* was associated with glycemic control and disease duration (r=0.4846, p=0.0043; r=0.7107, p=0.00174), whereas *WNT5A* and *GSK3B* were only correlated with glycemic control (r=0.5589, p=0.0037; r=0.4901, p=0.0051). Finally, Young’s Modulus was negatively correlated with *SOST* (r=-0.5675, p=0.0011), *AXIN2* (r=-0.5523, p=0.0042) and *SFRP5* (r=-0.4442, p=0.0437), while positively correlated with *LEF -1* (r=0.4116, p=0.0295) and *WNT10B* (r=0.6697, p=0.0001). These findings suggest that Wnt signaling, and AGEs could be the main determinants of bone fragility in T2D.

**Graphical abstract:** 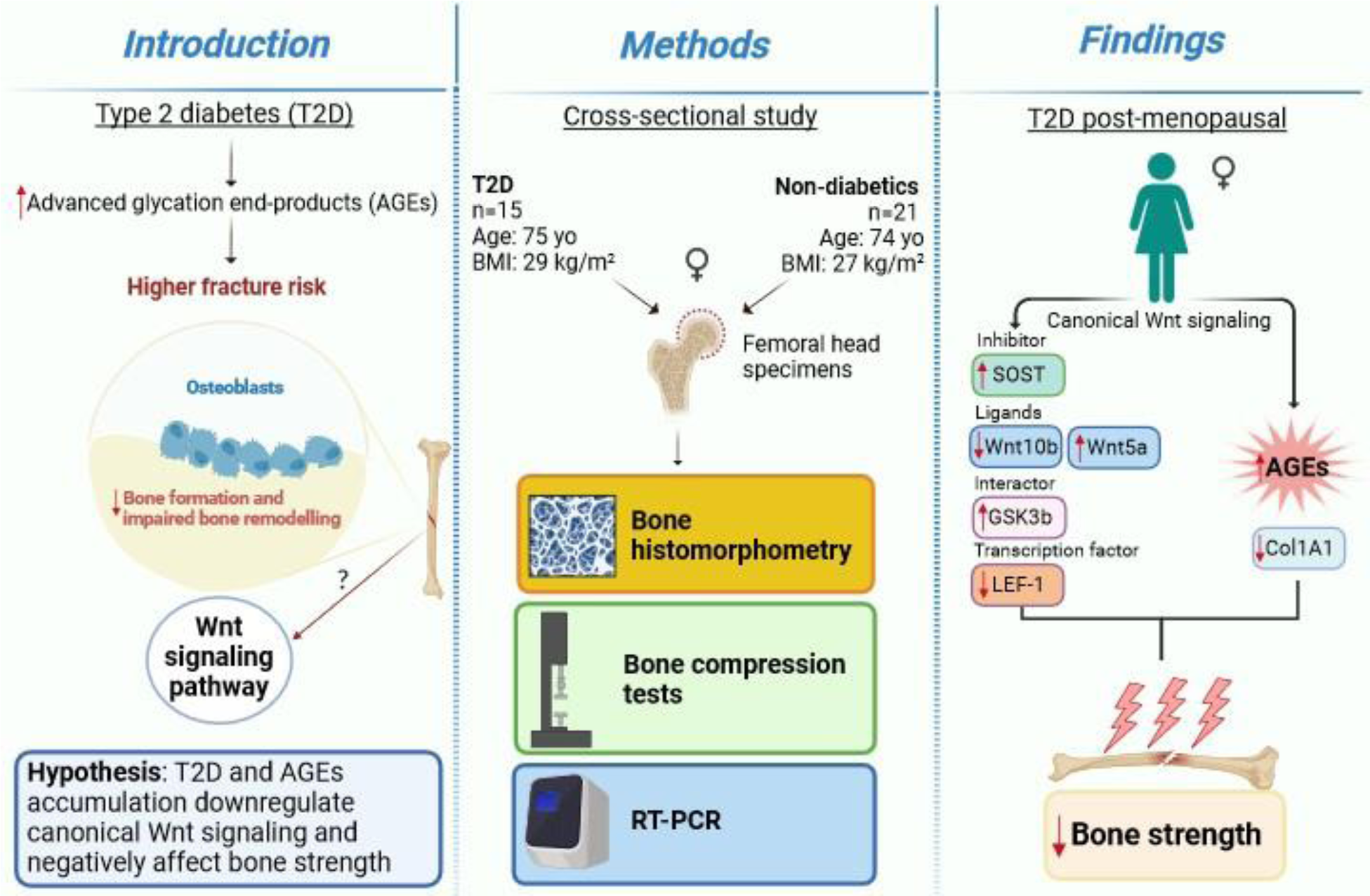

## Introduction

Type 2 diabetes (T2D) is a metabolic disease, with an increasing worldwide prevalence, characterized by chronic hyperglycemia and adverse effects on multiple organ systems, including bones [1]. Patients with T2D have an increased fracture risk, particularly at the hip, compared to individuals without diabetes. A recent meta-analysis reported that individuals with T2D have 1.27 relative risk (RR) of hip fracture compared to non-diabetic controls [2]. Fragility fractures in patients with T2D occur at normal or even higher bone mineral density compared to healthy subjects, implying compromised bone quality in diabetes. T2D is associated with a reduced bone turnover [3], as shown by lower serum levels of biochemical markers of bone formation, such as procollagen type1 amino-terminal propeptide (P1NP) and osteocalcin, and bone resorption, C-terminal cross-linked telopeptide (CTX) in diabetic patients compared to nondiabetic individuals [4–7]. Accordingly, dynamic bone histomorphometry of T2D postmenopausal women showed a lower bone formation rate, mineralizing surface, osteoid surface, and osteoblast surface[8]. Our group recently demonstrated that T2D is also associated with increased *SOST* and decreased *RUNX2* genes expression, compared to non-diabetic subjects[9]. Moreover, we have proved in a diabetic model that a sclerostin-resistant *Lrp5* mutation, associated with high bone mass, fully protected bone mass and strength even after prolonged hyperglycemia [10]. Sclerostin is a potent inhibitor of the canonical Wnt signaling pathway, a key pathway that regulates bone homeostasis [11].

Diabetes and chronic hyperglycemia are also characterized by increased advanced glycation end-products (AGEs) production and deposition [12]. AGEs may interfere with osteoblast differentiation, attachment to the bone matrix, function, and survival [13,14]. AGEs also alter bone collagen structure and reduce the intrinsic toughness of bone, thereby affecting bone material properties [9,15,16]. In this work,we hypothesized that T2D and AGEs accumulation downregulate Wnt canonical signaling and negatively affect bone strength.. Results confirmed that T2D downregulates Wnt beta/catenin signaling and reduces collagen mRNA levels and bone strength, in association with AGEs accumulation.

## Materials and Methods

### Study subjects

We enrolled a total of 36 postmenopausal women (15 with T2D and 21 non-diabetic controls) undergoing hip arthroplasty for osteoarthritis, consecutively screened to participate in this study between 2020 and 2022. Diabetes status was confirmed by the treating diabetes physician. Participants were diagnosed with diabetes when they had fasting plasma glucose (FPG) ≥126 mg/dl or 2-h plasma glucose (2-h PG) ≥200 mg/dl during a 75-g oral glucose tolerance test (OGTT); or hemoglobin A1c (HbA1c) ≥6.5% in accordance with the American Diabetes Association diagnostic criteria. Eligible participants were ≥60 years of age. Exclusion criteria were any diseases affecting bone or malignancy. Additionally, individuals treated with medications affecting bone metabolism such as estrogen, raloxifene, tamoxifen, bisphosphonates, teriparatide, denosumab, thiazolidinediones, glucocorticoids, anabolic steroids, and phenytoin, and those with hypercalcemia or hypocalcemia, hepatic or renal disorder, hypercortisolism, current alcohol or tobacco use were excluded. The study was approved by the Ethics Committee of the Campus Bio-Medico University of Rome and all participants provided written informed consent. All procedures were conducted in accordance with the Declaration of Helsinki.

### Specimen preparation

Femoral head specimens were obtained during hip arthroplasty. As described previously [9], trabecular bone specimens were collected fresh and washed multiple times in sterile PBS until the supernatant was clear of blood. Bone samples were stored at – 80 °C until analysis.

### Bone histomorphometry

Trabecular bone from femur heads werefixed in 10% neutral buffered formalin for 24 h prior to storage in 70% ethanol. Tissues were embedded in methylmethacrylate and sectioned sagittally by the Washington University Musculoskeletal Histology and Morphometry Core. Sections were stained with Goldner’s trichrome. Then, a rectangular region of interest containing trabecular bone was chosen below the cartilage-lined joint surface and primary spongiosa. This region had an average dimension of 45 mm^2^. Tissue processing artifacts, such as folding and edges, were excluded from the ROI. A threshold was chosen using the Bioquant Osteo software to automatically select trabeculae and measure bone volume. Finally, Osteoid was highlighted in the software and quantified semi-automatically using a threshold and correcting with the brush tool. Unstained and TRAP-stained (Sigma) slides were imaged at ×20 high resolution using a NanoZoomer 2.0 with bright field and FITC/TRITC (Hamamatsu Photonics). Images were then analyzed via Bioquant Osteo software according to the manufacturer’s instructions and published standards (v18.2.6, Bioquant Image Analysis Corp., Nashville, TN).

### Bone compression tests

We used cylindrical bone specimens of trabecular core (with a diameter of 10 mm and a length of 20 mm) from 10 T2D and 21 non-diabetic subjects to measure bone mechanical parameters (Young’s modulus, ultimate strength and yield strength), as previously described [9].

### RNA extraction and gene expression by RT-PCR

Total RNA from trabecular bone samples was extracted using TRIzol (Invitrogen) following the manufacturer’s instructions. The concentration and purity of the extracted RNA were assessed spectrophotometrically (TECAN, InfiniteM200PRO), and only samples with 260/280 absorbance ratio between 1.8 and 2 were used for reverse transcription using High-Capacity cDNA Reverse Transcription Kit (Applied Biosystems, Carlsbad, CA) according to the manufacturer’s recommendations. Transcription products were amplified using TaqMan real-time PCR (Applied Biosystems, Carlsbad, CA) and a standard protocol (95°C for 10 minutes; 40 cycles of 95°C for 15 seconds and 60°C for 1 minute; followed by 95°C for 15 seconds, 60°C for 15 seconds, and 95°C for 15 seconds). β-Actin expression was used as an internal control (housekeeping gene). Relative expression levels of Sclerostin (*SOST*), Dickkopf-1 (*DKK-1*), Wnt ligands (*WNT5A* and *WNT10B*), T-cell factor/ lymphoid enhancer factor 1 (*LEF-1*), collagen type I alpha 1 chain (*COL1A1*), glycogen synthase kinase 3 beta (*GSK3B*), axis inhibition protein 2 (*AXIN2*), beta-catenin (*BETA-CATENIN*) and secreted frizzled-related protein 5 (*SFRP5*) were calculated using the 2^-ΔCt^ method.

### Statistical analysis

Data were analyzed using GraphPad Prism 9.0 (GraphPad Software, San Diego, CA). Patients’ characteristics were described using means and standard deviations or medians and 25th-75th percentiles, as appropriate, and percentages. Group data are presented in boxplots with median and interquartile range; whiskers represent maximum and minimum values. We assessed data for normality and Mann-Whitney test was used to compare variables between groups. Data were analyzed using nonparametric Spearman correlation analysis and the correlation coefficients (r) were used to assess the relationship between variables. We used Grubbs’ Test to assess and exclude outliers.

## RESULTS

### Subject characteristics

Clinical characteristics of study subjects are presented in Table 1. T2D and non-diabetic subjects did not differ in age, BMI and menopausal age. As expected, fasting glucose was significantly higher in T2D compared to non-diabetic subjects [112.00 mg/dl (104.0-130.0 ) mg/dl, *vs.* 94.00 (87.2-106.3), respectively; p=0.009]. Median HbA1c was determined in all T2D subjects within three months before surgery [(6.95% (6.37-7.37)]. Median disease duration in T2D subjects was [14.50 years (7.25-19.25)]. Diabetes medications included monotherapy with metformin (n=12) and combination therapy with metformin plus insulin and glinide (n=3). There were no differences in serum calcium, eGFR (CKD-EPI equation) and serum blood urea nitrogen.

**Table 1.** Results were analyzed using unpaired T-test with Welch’s correction. Results are presented as median and percentiles (25th and 75th).

|  | **T2D subjects**<br>**(n=15)** | **Non-diabetic**<br>**subjects**<br>**(n=21)** | **P value** |
| --- | --- | --- | --- |
| Age (years) | 73.00 (67.00-80.00) | 73.00 (68.50-79.00) | 0.644 |
| BMI (kg/m<sup>2</sup>) | 30.81 (24.44-34.00) | 25.00 (24.00-31.50) | 0.117 |
| Menopausal age (years) | 50.00 (42.50-52.75) | 52.00 (48.00-53.00) | 0.344 |
| Fasting glucose levels (mg/dl) | 112.00 (104.00-130.0) | 94.00 (87.25-106.3) | **0.009 |
| Disease duration (years) | 14.50 (7.25-19.25) | - | - |
| HbA1c (%) | 6.95 (6.37-7.37) | - | - |
| Serum calcium (mg/dl) | 9.05 (8.800-9.550) | 9.15 (9.000-9.550) | 0.535 |
| eGFR (mL/min/1.73 m<sup>2</sup>) | 78,30 (59.90-91.10) | 75.60 (61.35-88.55) | 0.356 |
| Serum blood urea nitrogen (mg/dl) | 42.00 (36.00-53.00) | 37.00 (31.75-46.50) | 0.235 |

### Bone Histomorphometry

Bone samples of 8 T2D and 9 non-diabetic subjects were used for histomorphometry analysis. We found no significant differences in BV/TV and osteoid volume, while mineralized volume/total volume (Md.V/TV) trended lower in T2D subjects relative to controls [0.249% (0.156-0.336) vs 0.352% (0.269-0.454); p=0.053] (Table 2).

**Table 2.** Results were analyzed using unpaired T-test with Welch’s correction and are presented as median and percentiles (25th and 75th).

|  | T2D subjects<br>(n=8) | Non-diabetic<br>subjects<br>(n=9) | P value |
| --- | --- | --- | --- |
| BV/TV (%) | 0.248 (0.157-0.407) | 0.358 (0.271-0.456) | 0.120 |
| Md.V/BV (%) | 0.994(0.984-0.998) | 0.995 (0.985-0.997) | 0.998 |
| Md.V/TV (%) | 0.249 (0.156-0.366) | 0.352 (0.269-0.454) | 0.053 |
| OV/BV (%) | 0.009 (0.002-0.009) | 0.004 (0.002-0.015) | 0.704 |
| OV/TV (%) | 0.001 (0.0002-0.0058) | 0.001 (0.0007-0.0056) | 0.896 |
| OS/BS (%) | 0.026 (0.022-0.161) | 0.035 (0.009-0.117) | 0.525 |

### Bone compression tests

Young’s modulus was lower in T2D compared to non-diabetic subjects [21.6 MPa (13.46-30.10) vs. 76.24 MPa (26.81-132.9); p=0.0025), while ultimate strength and yield strength were not different between the two groups (Table 3).

**Table 3.** Results were analyzed using unpaired T-test with Welch’s correction and are presented as median and percentiles (25th and 75th).

|  | T2D subjects<br>(n=10) | Non-diabetic<br>subjects<br>(n=21) | P value |
| --- | --- | --- | --- |
| Young's modulus (MPa) | 21.60 (13.46-30.10) | 76.24 (26.81-132.9) | 0.002 |
| Ultimate strength (MPa) | 3.015 (2.150-13,86) | 7.240 (3.150-8.898) | 0.914 |
| Yield strength (MPa) | 2.525 (1.943-6.393) | 6.150 (3.115-7.423) | 0.159 |

### Gene Expression

*SOST* mRNA was significantly higher in T2D than in non-diabetic subjects (Fig. 1A, p<0.0001), whereas there was no difference in *DKK1* gene expression between the two groups (Fig. 1B). Of note, *SOST* mRNA transcript was very low in the majority of non-diabetic subjects (Fig. 1A). *LEF-1* (Fig. 1C, p=0.0136), *WNT10B (Fig. 1D*, p=0.0302) and *COL1A1* (Fig.1F, p=0.0482) mRNA transcripts were significantly lower in T2D compared to non-diabetic subjects. Conversely, *WNT5A* was higher in T2D relative to non-diabetics (Fig. 1E, p=0.0232). Moreover, *GSK3B* was significantly increased in T2D compared to non-diabetic subjects (Fig.1G, p=0.0456), but we did not find any significant difference in gene expression of *AXIN2*, *BETA-CATENIN* and *SFRP5* (Fig. 1H-J) between our groups.

**Figure 1.**
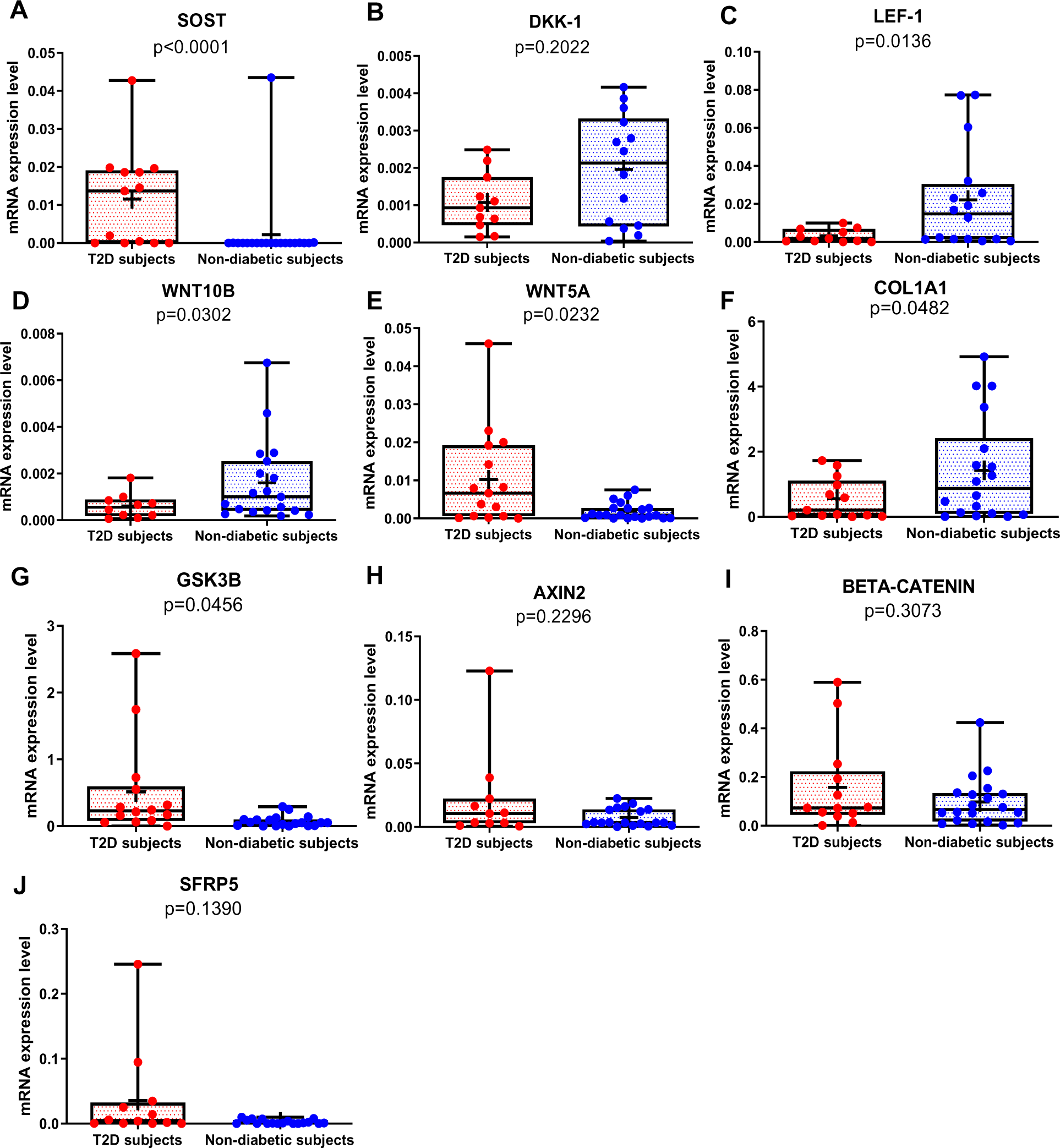
Gene expression analysis in trabecular bone samples. (A) SOST mRNA levels resulted higher in T2D subjects versus Nondiabetic subjects (p<0.0001). (B) DKK-1 mRNA expression level was not different between groups (p=0.2022). (C) LEF-1 mRNA levels resulted lower in T2D subjects versus nondiabetics subjects (p=0.0136). (D) WNT10B mRNA expression level was lower in T2D subjects versus nondiabetic subjects (p=0.0302). (E) WNT5A mRNA resulted higher in T2D subjects versus nondiabetics subjects (p=0.0232). (F) COL1A1 mRNA levels resulted lower in T2D subjects versus Nondiabetic subjects (p=0.0482). (G) GSK3B mRNA levels resulted higher in T2D subjects versus Nondiabetic subjects (p=0.0456). (H-J) AXIN2, BETA-CATENIN, SFRP5 mRNA levels were not different between groups (p=0.2296, p=0.3073; p=0.1390). Data are expressed as fold changes over beta-actinMedians and interquartile ranges, differences between non-diabetics and T2D subjects were analyzed using Mann-Whitney test.

### Correlation analysis of Wnt target genes, AGEs and glycemic control

As shown in figure 2, AGEs were inversely correlated with *LEF-1* (Fig. 2A, p=0.0255) and *COL1A1* mRNA abundance (Fig. 2B, p=0.0004), whereas they were positively correlated with *SOST* (Fig. 2C, p<0.0001) and *WNT5A* mRNA (Fig. 2D, p=0.0322). There was no correlation between AGEs content and *WNT10B* (Fig. 2E; p=0.1938) or *DKK1* gene expression (Fig. 2F; p=0.9349). Likewise, we did not find any significant correlation between *LEF-1*, *WNT5A*, *WNT10B*, *DKK-1*, *COL1A1* expression in bone and glycemic control in T2D individuals (Supplemental Figure 1A-D). However, there were positive correlations between *SOST* and fasting glucose levels (Fig. 3A, p=0.0043), *SOST* and disease duration (Fig. 3B, p=0.00174), *WNT5A, GSK3B* and fasting glucose levels (Fig. 3C, p=0.0037; Fig. 3D, p=0.0051).

**Figure 2.**
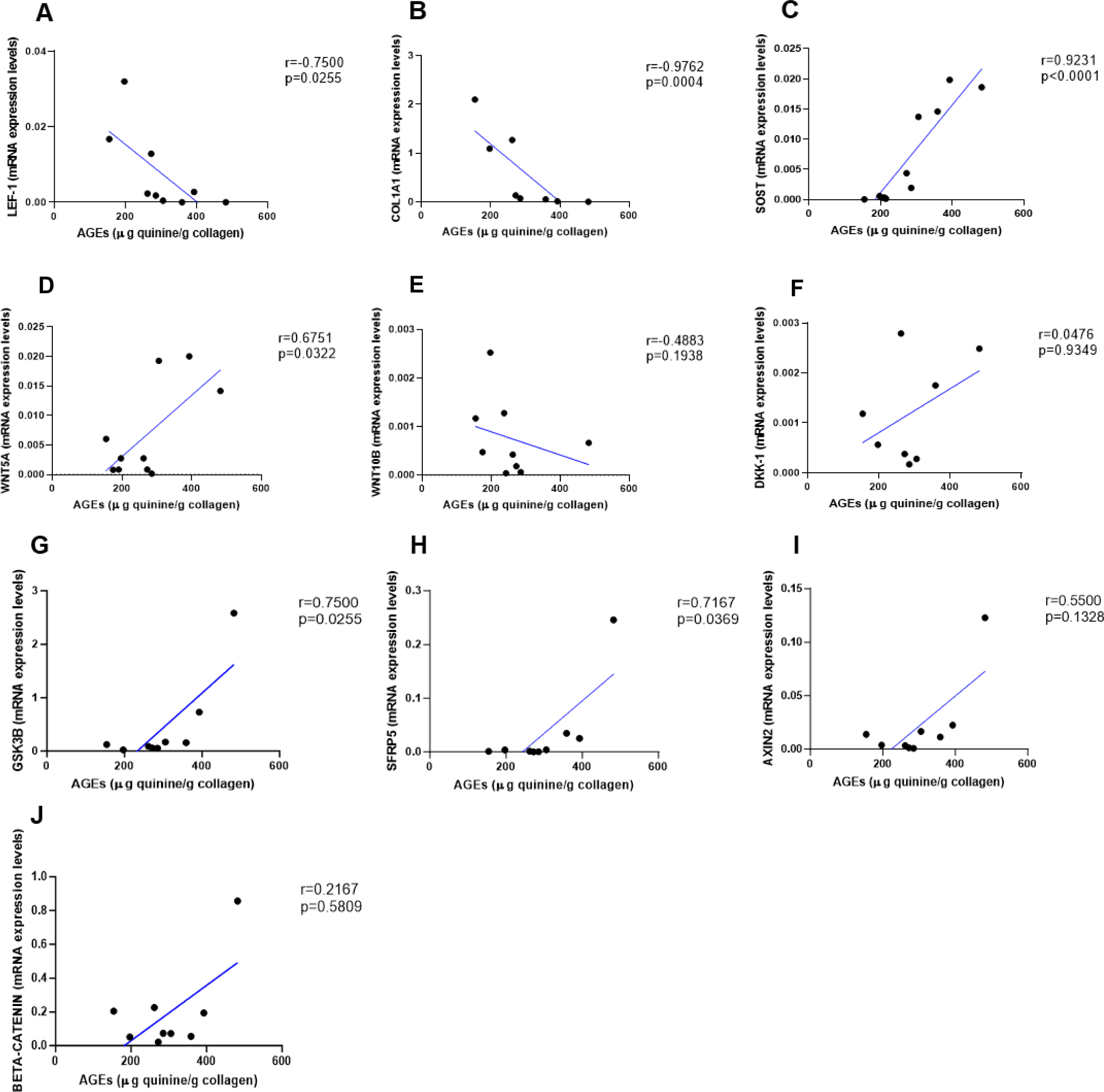
Relationship between AGEs (µg quinine/g collagen) bone content and mRNA level of the Wnt signaling key genes in T2D and non-diabetic subjects. (A) LEF-1 negatively correlated with AGEs (r=-0.7500; p=0.0255). (B) COL1A1 negatively correlated with AGEs (r=-0.9762; p=0.0004). (C) SOST mRNA level expression positively correlated with AGEs (r=0.9231; p<0.0001). (D) WNT5A mRNA expression level positively correlated with AGEs (r=0.6751; p=0.0322). (E) WNT10B mRNA expression level was not correlated with AGEs (r=-0.4883; p=0.1938). (F) DKK1 mRNA expression level was not correlated with AGEs (r=0.0476; p=0.9349). (G) GSK3B mRNA expression level was positively correlated with AGEs (r=0.7500; p=0.0255). (H) SFRP5 mRNA expression level was positively correlated with AGEs (r=0.7167; p=0.0369). (I) AXIN2 and (J) SFRP5 mRNA expression levels were not correlated with AGEs (r=0.5500, p=0.1328; r=0.2167, p=0.5809). Data were analyzed using nonparametric Spearman correlation analysis and r represents the correlation coefficient.

**Figure 3.**
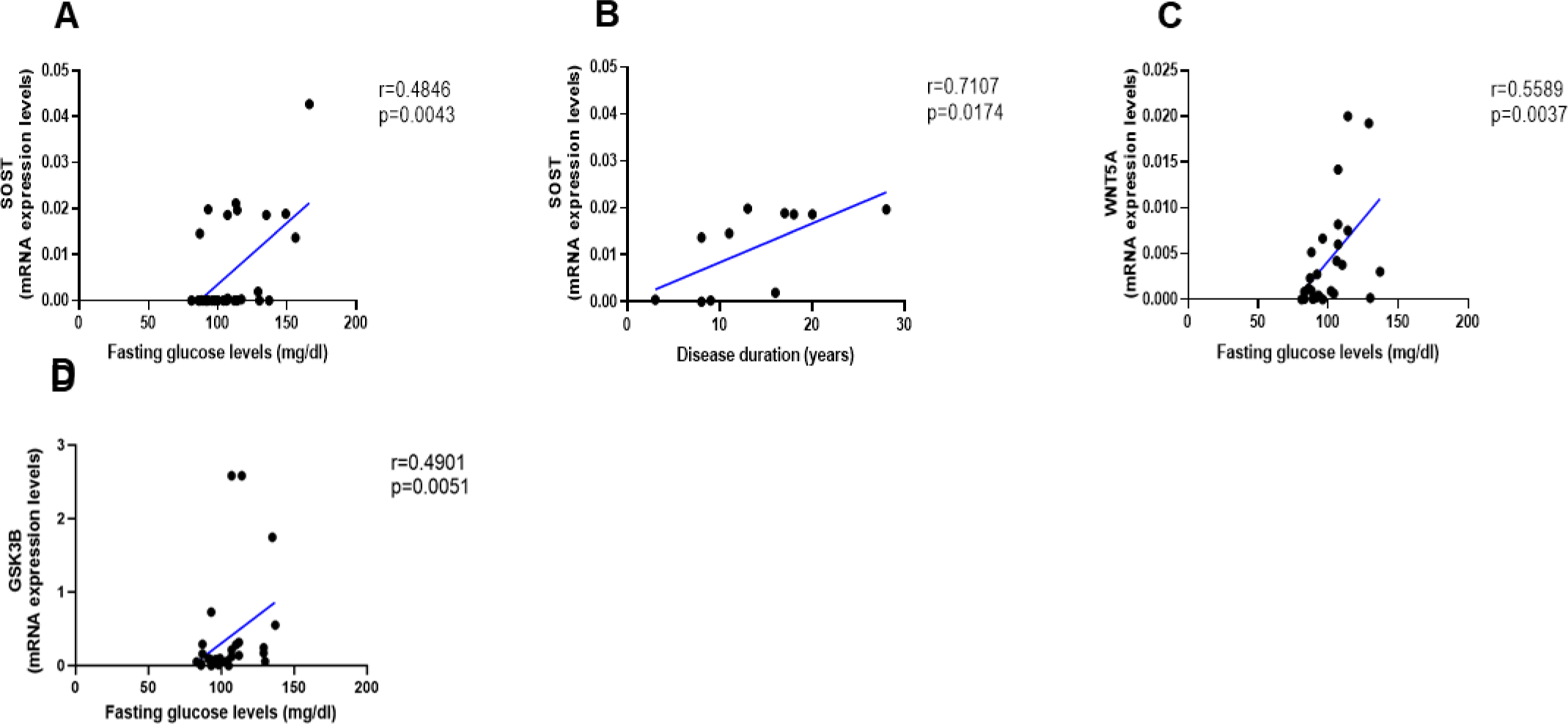
Relationship between fasting glucose levels (mg/dl) and disease duration with SOST and WNT5A mRNA levels. (A) SOST positively correlated with fasting glucose levels (r=0.4846; p=0.0043). (B) SOST positively correlated with disease duration (r=0.7107; p=0.0174). (C) WNT5A positively correlated with fasting glucose levels (r=0.5589; p=0.0037). (D) GSK3B positively correlated with fasting glucose levels (r=0.4901; p=0.0051). Data were analyzed using nonparametric Spearman correlation analysis and r represents the correlation coefficient.

### Correlation analysis of Wnt target genes and bone mechanical parameters

As shown in Figure 4, Young’s Modulus was negatively correlated with *SOST* (Fig.4A, p=0.0011), *AXIN2* (Fig. 4D, p=0.0042) and *SFRP5* (Fig 4F, p=0.0437), while positively correlated with *LEF -1* (Fig. 4B, p=0.0295) and *WNT10B* (Fig. 4C, p=0.0001). Ultimate strength was associated with *WNT10B* (Fig. 4F, p=0.0054) and negatively correlated with *AXIN2* (Fig. 4G, p=0.0472). Finally, yield strength was associated with *LEF-1* (Fig. 4H, p=0.0495) and *WNT10B* (Fig. 4I, p=0.0020) and negatively correlated with *GSK3B* (Fig. 4J, p=0.0245), AXIN2 (Fig. 4K, p=0.0319), and *SFRP5* (Fig. 4L, p=0.0422). Non-significant correlations are reported in Supplementary figure 2 (A-Q).

**Figure 4.**
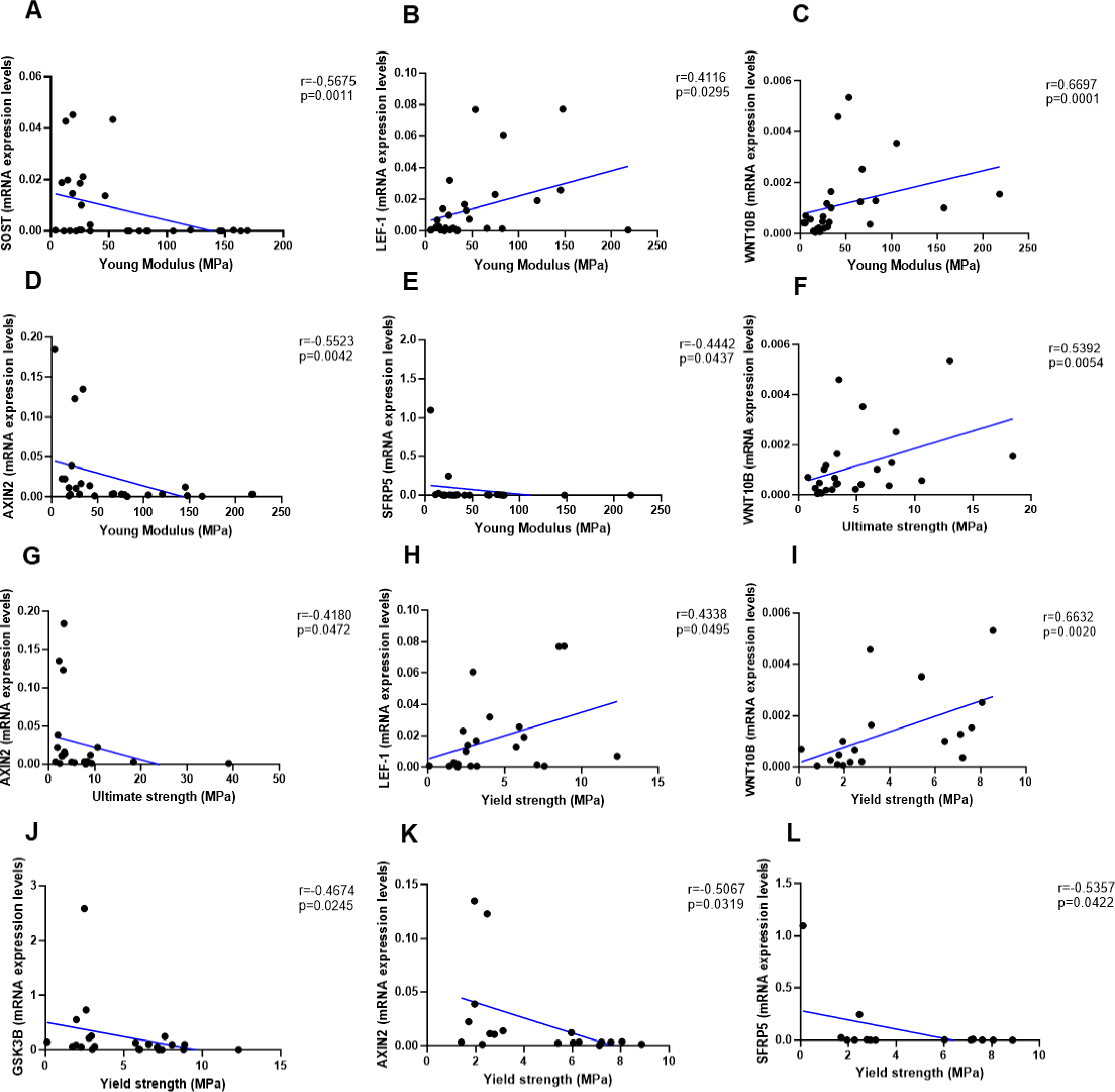
Relationship between Young Modulus (MPa), Ultimate strength (MPa) and Yield strength (MPa) with mRNA levels of the Wnt signaling key genes in T2D and non-diabetic subjects. (A) SOST negatively correlated with Young Modulus (MPa); (r=-0.5675; p=0.0011). (B) LEF-1 positively correlated with Young Modulus (MPa); (r=0.4116; p=0.0295). (C) WNT10B positively correlated with Young Modulus (MPa); (r=0.6697; p=0.0001). (D) AXIN2 negatively correlated with Young Modulus (MPa); (r=-0.5523; p=0.0042). (E) BETA-CATENIN negatively correlated with Young Modulus (MPa); (r=-0.5244; p=0.0050). (F) SFRP5 negatively correlated with Young Modulus (MPa); (r=-0.4442; p=0.0437). (G) WNT10B positively correlated with Ultimate strength (MPa); (r=0.5392; p=0.0054). (H) AXIN2 negatively correlated with Ultimate strength (MPa); (r=-0.4180; p=0.0472). (I) BETA-CATENIN negatively correlated with Ultimate strength (MPa); (r=-0.5528; p=0.0034). (J) LEF-1 positively correlated with Yield strength (MPa); (r=0.4338; p=0.0495). (K) WNT10B positively correlated with Yield strength (MPa); (r=0.6632; p=0.0020). (L) GSK3B negatively correlated with Yield strength (MPa); (r=-0.4674; p=0.0245). (M) AXIN2 negatively correlated with Yield strength (MPa); (r=-0.5067; p=0.0319). (N) BETA-CATENIN negatively correlated with Yield strength (MPa); (r=-0.5491; p=0.0149). (O) SFRP5 negatively correlated with Yield strength (MPa); (r=-0.5357; p=0.0422). Data were analyzed using nonparametric Spearman correlation analysis and r represents the correlation coefficient.

## Discussion

We show that key components of the Wnt/beta-catenin signaling are abnormally expressed in the bone of postmenopausal women with T2D and they are associated with AGEs and reduced bone strength. *LEF-1*, a transcription factor that mediates responses to Wnt signal and Wnt target genes itself, and *WNT10B*, an endogenous regulator of Wnt/β-catenin signaling and skeletal progenitor cell fate, are both downregulated in bone of postmenopausal women with T2D. Consistently, in this group, the expression of the Wnt inhibitor, *SOST* is increased, suggesting suppression of Wnt/β-catenin signaling. Interestingly, our data suggest that sclerostin expression is very low in healthy postmenopausal women not affected by osteoporosis. Moreover, we reported an increase in the expression level of bone *GSK3B,* in line with downregulated Wnt/β-catenin signaling in T2D. Our data also show that the expression of *WNT5A*, a non-canonical ligand linked to inhibition of Wnt/beta-catenin signaling was increased, whereas *COL1A1* was decreased. These findings are consistent with reduced bone formation and suppression of Wnt signaling in T2D. We have previously reported upregulation of *SOST* and downregulation of *RUNX2* mRNA in another cohort of postmenopausal women with T2D [9]. Of note, the cohort of T2D subjects studied here had glycated hemoglobin within therapeutic targets, implying that the changes in gene transcription we identified persist in T2D bone despite good glycemic control.

High circulating sclerostin has been reported in diabetes [17,18], and increased sclerostin is associated with fragility fractures [19]. Aside from confirming higher *SOST* expression, we also show that other Wnt/β-catenin osteogenic ligands are abnormally regulated in the bone of T2D postmenopausal women. *WNT10B* is a positive regulator of bone mass; transgenic overexpression in mice results in increased bone mass and strength [20], whereas genetic ablation of *WNT10B* is characterized by reduced bone mass [21,22], and decreased number and function of osteoblasts [21]. More to the point, *WNT10B* expression is reduced in the bone of diabetic mice [23]. Therefore, the reduced *WNT10B* in human bone we found in the present study further supports the hypothesis of reduced bone formation in T2D. Accordingly, *LEF-1* gene expression was also downregulated confirming that Wnt/beta-catenin pathway is decreased in T2D. Importantly, the overexpression of *LEF-1* induces the expression of osteoblast differentiation genes (osteocalcin and *COL1A1*) in differentiating osteoblasts [24]. In fact, in this study we also demonstrated that a downregulation of *LEF-1* in T2D bone goes along with a downregulation of *COL1A1*, strengthen data of a reduced production of bone matrix most likely as the result of reduced osteoblasts synthetic activity in diabetes [8,25]. Reduced *RUNX2* in T2D postmenopausal women also confirms previous findings [9] and further supports the notion of reduced osteoblast differentiation or function in diabetes. On the other hand, the contribution of upregulated *WNT5A* in diabetic bone is more complex. *WNT5A* regulates Wnt/beta-catenin signaling depending on the receptor availability [26]. Non-canonical *WNT5A* activates β-catenin-independent signaling, including the Wnt/Ca^++^[27] and planar cell polarity pathways [28].

Heterozygous Wnt5a null mice have low bone mass with impaired osteoblast and osteoclast differentiation [29]. Wnt5a inhibits Wnt3a protein by downregulating beta/catenin-induced reporter gene expression [26]. In line with these findings, we showed that there was an increased gene expression of *WNT5A* in bone of T2D postmenopausal women, confirming a downregulated Wnt/beta-catenin signaling and impaired osteoblasts function. Moreover, *GSK3B* is a widely expressed serine/threonine kinase involved in multiple pathways regulating immune cell activation and glucose metabolism. Preclinical studies reported that *GSK3B* is a negative regulator of Wnt/beta-catenin signaling and bone metabolism [30,31], and its increase is associated with T2D and alterations in insulin secretion and sensitivity [32,33]. Our data, confirmed that *GSK3B* is increased in T2D postmenopausal women and it is associated with reduced yield strength. In fact, we also showed an impaired bone mechanical plasticity in T2D, in line with other studies showing a reduced bone strength [16,34,35]. In addition, this study reported significant correlations of bone mechanical parameters and Wnt target genes, which might reflect the biological effect of downregulated Wnt signaling and AGEs accumulation on bone mechanical properties in diabetes.

We have previously shown that AGEs content is higher in T2D bone compared to non-diabetic bone, even in patients with well-controlled T2D [9]. Here we show that AGEs accumulation is positively correlated with *SOST*, *WNT5A* and *GSK3B* gene expression, and negatively correlated with *LEF-1*, *WNT10B*, and *COL1A1* mRNA. These findings are consistent with the hypothesis that AGEs accumulation is associated with impaired Wnt signaling and low bone turnover in T2D. We did not find any abnormalities in histomorphometric parameters in our subjects with T2D, consistent with our previous report [9]. Reduced osteoid thickness and osteoblast number were reported in premenopausal T2D women with poor glycemic control compared to non-diabetic subjects but not in the group with good glycemic control [36]. Therefore, good glycemic control appears to prevent or rescue any changes in static histologic parameters of bone turnover that might be caused by uncontrolled diabetes.

Our study has some limitations. One is the cross-sectional design; another one is the relatively small number of T2D subjects enrolled. Moreover, we measured the mRNA abundance of the genes of interest, and we cannot assume that the differences we found reflect differences in protein abundance. Although osteoarthritis may affect some of the genes we studied [37], all study subjects were affected by variable degree of osteoarthritis, and the effect of such potential confounder is not likely to be different between T2D and control subjects. Finally, we did not use the tetracycline double-labeled technique to investigate dynamic bone parameters.

The main strength of our study is that this study is the first to explore the association of AGEs on Wnt pathway in postmenopausal T2D women. Moreover, we measured the expression of several Wnt genes directly on bone samples of postmenopausal T2D women.

In conclusion, our data show that, despite good glycemic control, T2D decreases expression of *COL1A1* and Wnt genes that regulate bone turnover, in association with increased AGEs content and reduced bone strength. These results may help understand the mechanisms underlying bone fragility in T2D.

## Data Availability

All data are available upon request to the corresponding authors

## Acknowledgements

This work was supported by an internal Grant of Campus Bio-Medico University of Rome.

## Author Contributions

Conceptualization, N.N, R.P., M.M and R.S.; methodology, G.L., F.C., M.F., V.V., F.T., N.P., A.B., S.T..; software, G.L., C.P., A.B., and E.S.; validation, N.N. and G.L.; formal analysis, N.N. and G.L.; investigation, N.N., R.S., M.M. and R.P.; resources, N.N, G.V., R.P. M.M., and R.S.; data curation, G.L. and N.P..; writing—original draft preparation, G.L., M.F., N.P., R.C. and N.N.; writing—review and editing, G.L., N.N., C.P., N.P., F.Z., E.S., R.S. and R.C.; visualization, N.N. and R.S..; supervision, N.N., R.P., and M.M.; project administration, N.N. and G.L.; funding acquisition, N.N. and R.S. All authors have read and agreed to the published version of the manuscript.

## Legend of supplementary figures

**Supplementary figure 1.**
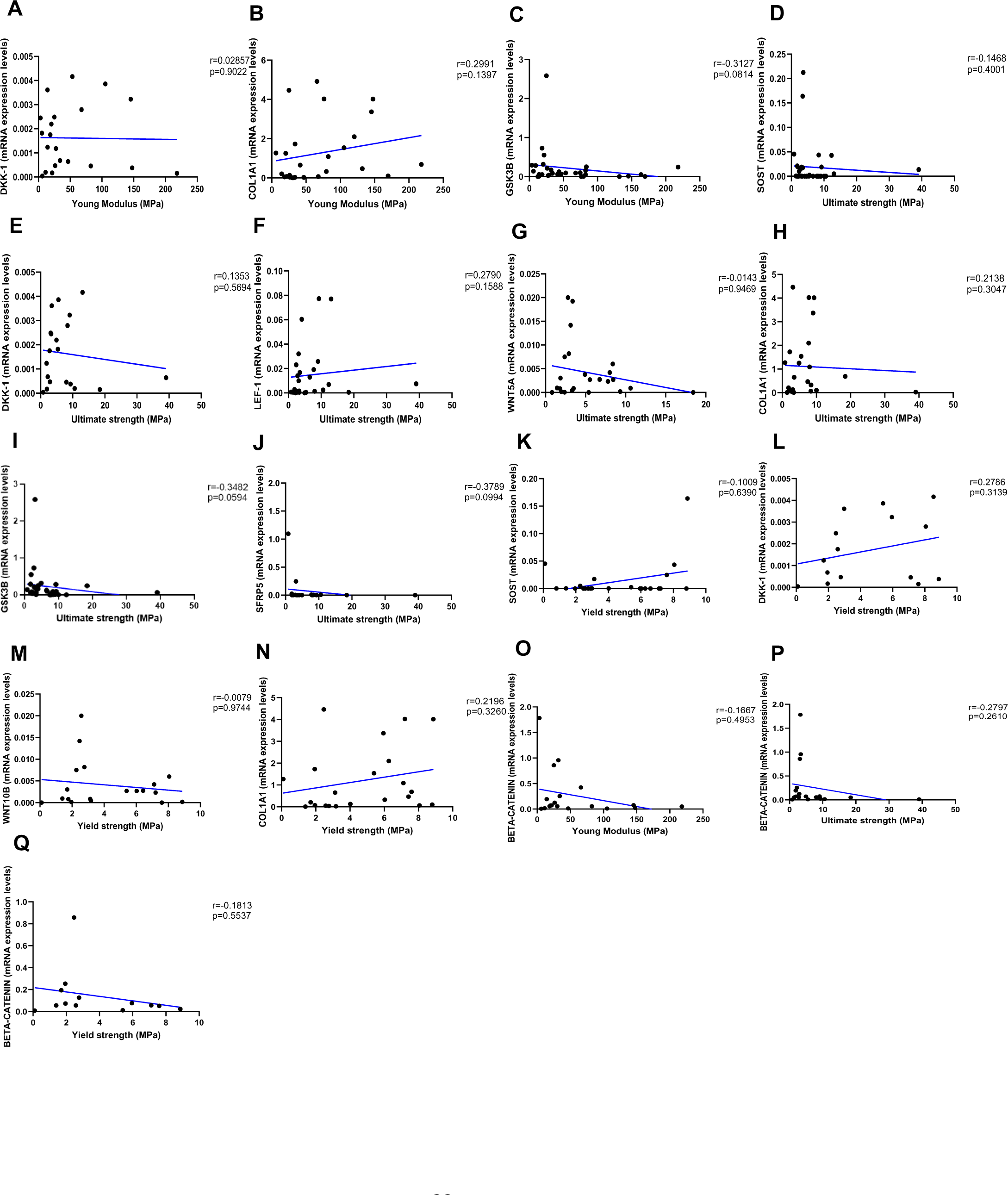
Relationship between fasting glucose levels (mg/dl) and LEF-1, WNT5A, WNT10B, DKK-1, COL1A1 mRNA levels. (A-E) Data showed negative correlations between fasting glucose levels (mg/dl) and (A) LEF-1 (r=-0.3649; p=0.0613), (B) WNT10B (r=-0.0041; p=0.9863), (C) COL1A1 (r=-0.1157; p=0.5354), (D) DKK-1 (r=-0.0947; p=0.6522) mRNA levels. Data showed positive correlations between fasting glucose levels (mg/dl) with (E) AXIN2 (r=0.0993; p=0.6442), (F) BETA-CATENIN (r=0.2371; p=0.1991) and (G) SFRP5 (r=0.3767; p=0.0696). Data were analyzed using nonparametric Spearman correlation analysis and r represents the correlation coefficient.

**Supplementary figure 2.**
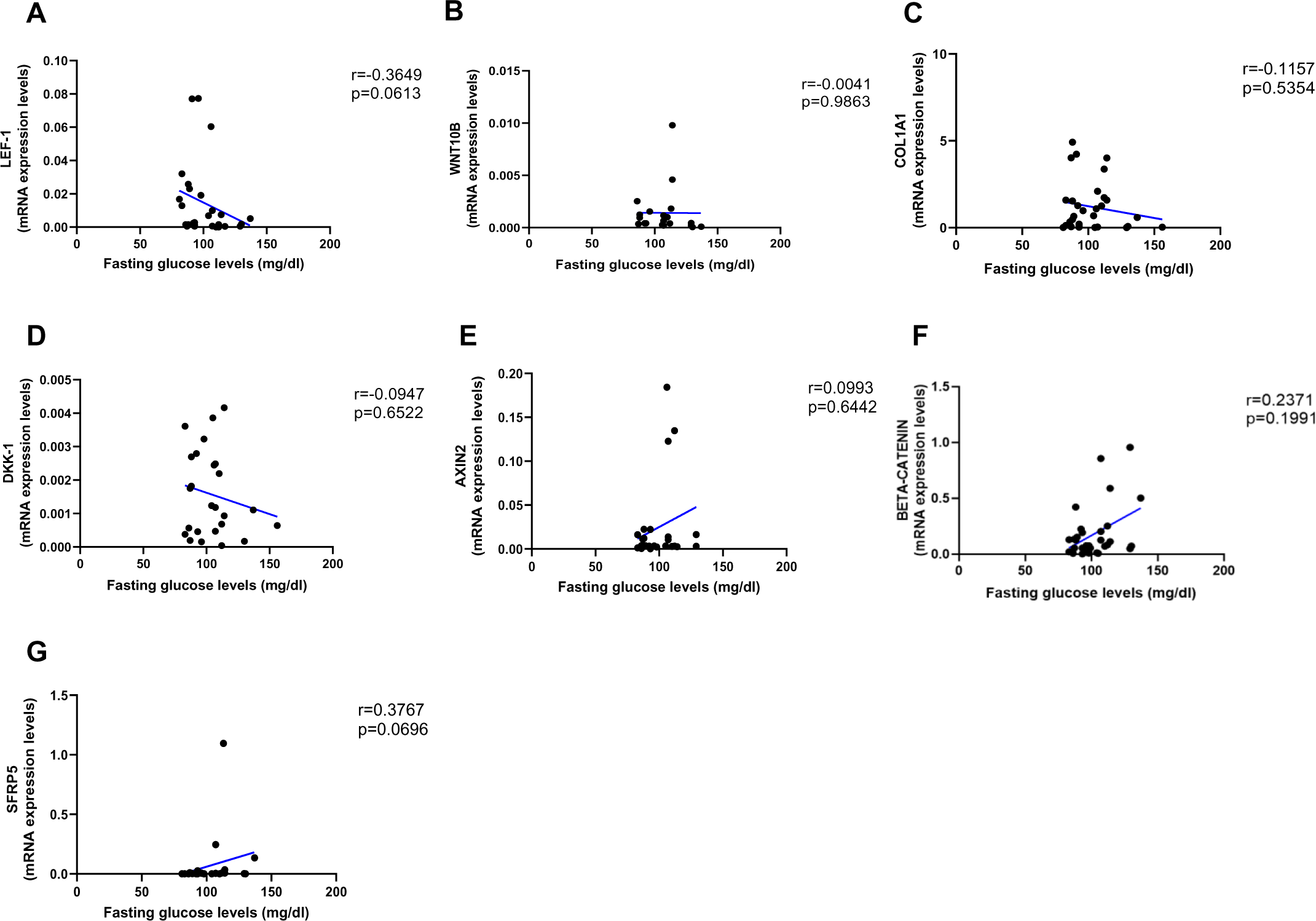
Relationship between Young Modulus (MPa), Ultimate strength (MPa) and Yield strength (MPa) with mRNA levels of the Wnt signaling genes in T2D and non-diabetic subjects. (A) DKK-1 positively correlated with Young Modulus (MPa); (r=0.02857; p=0.9022). (B) COL1A1 positively correlated with Young Modulus (MPa); (r=0.2991; p=0.1397). (C) GSK3B negatively correlated with Young Modulus (MPa); (r=-0.3127; p=0.0814). (D) SOST negatively correlated with Ultimate strength (MPa); (r=-0.1468; p=0.4001). (E) DKK-1 negatively correlated with Ultimate strength (MPa); (r=0.1353; p=0.5694). (F) LEF-1 positively correlated with Ultimate strength (MPa); (r=0.2790; p=0.1588). (G) WNT5A negatively correlated with Ultimate strength (MPa); (r=-0.0143; p=0.9469). (H) COL1A1 positively correlated with Ultimate strength (MPa); (r=0.2138; p=0.3047). (I) GSK3B negatively correlated with Ultimate strength (MPa); (r=-0.3482; p=0.0594). (J) SFPR5 negatively correlated with Ultimate strength (MPa); (r=-0.3789; p=0.0994). (K) SOST positively correlated with Yield strength (MPa); (r=0.1009; p=0.6390). (L) DKK-1 positively correlated with Yield strength (MPa); (r=0.2786; p=0.3139). (M) WNT10B negatively correlated with Yield strength (MPa); (r=-0.0079; p=0.9744). (N) COL1A1 positively correlated with Yield strength (MPa); (r=0.2196; p=0.3260). (O) BETA-CATENIN negatively correlated with Young Modulus strength (MPa); (r=-0.1667; p=0.4953). (P) BETA-CATENIN negatively correlated with Ultimate strength (MPa); (r=-0.2797; p=0.2610). (Q) BETA-CATENIN negatively correlated with Yield strength (MPa); (r=-0.1813; p=0.5537). Data were analyzed using nonparametric Spearman correlation analysis and r represents the correlation coefficient.

