## Supplementary figures and images for "Bone canonical Wnt signaling is downregulated in type 2 diabetes and associates with higher Advanced Glycation End-products (AGEs) content and reduced bone strength"

### Supplemental figure 1

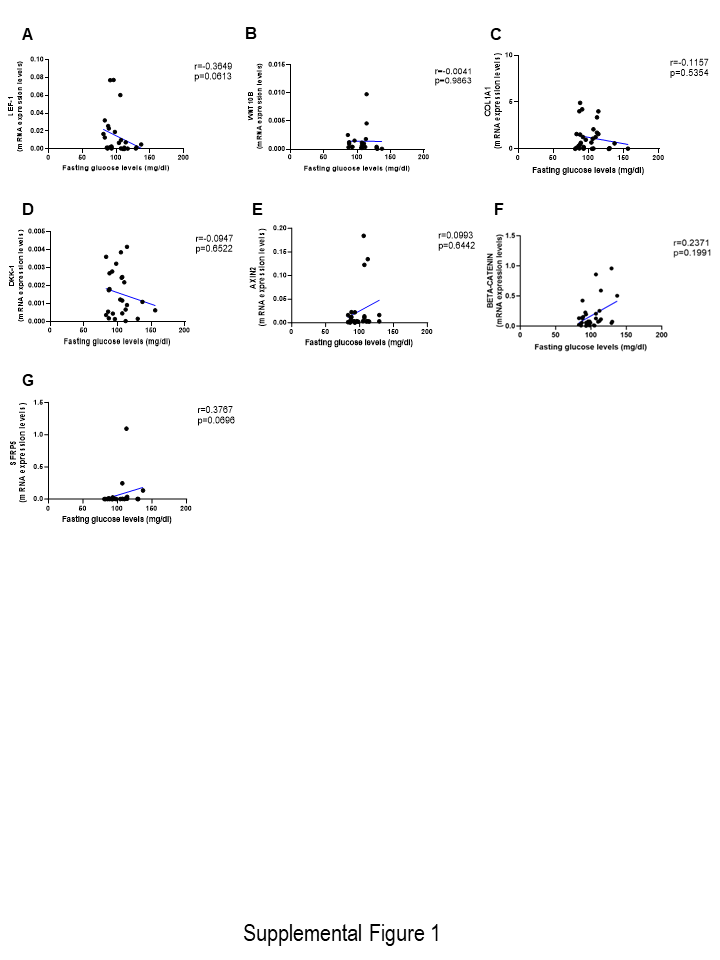

### Supplemental figure 2

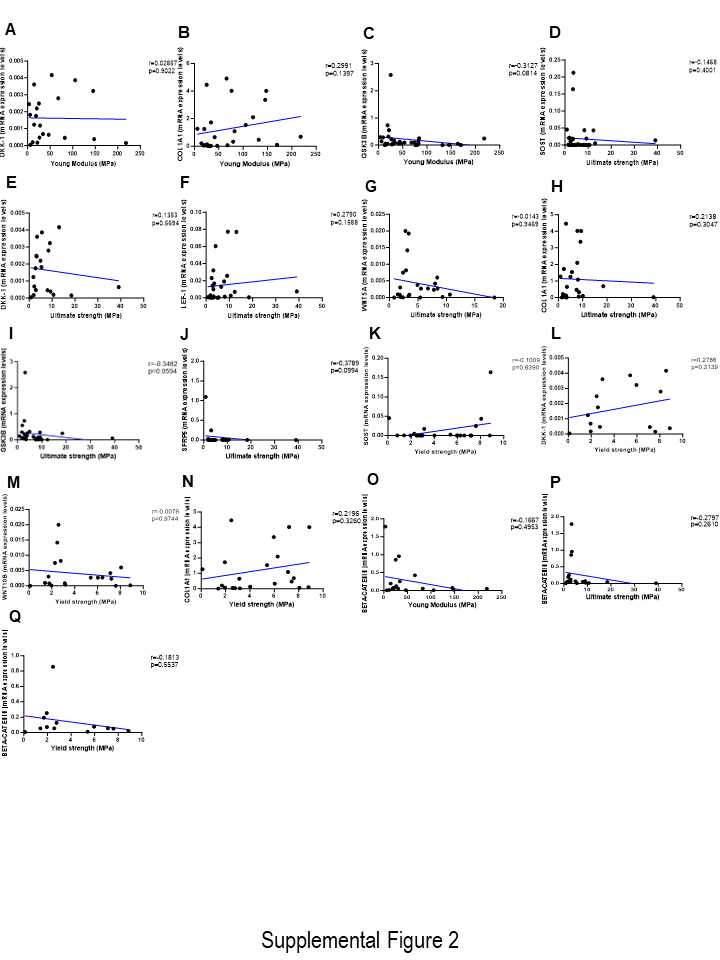
